# Epidemiological characteristics of 1212 COVID-19 patients in Henan, China

**DOI:** 10.1101/2020.02.21.20026112

**Authors:** Pei Wang, Jun-an Lu, Yanyu Jin, Mengfan Zhu, Lingling Wang, Shunjie Chen

**Affiliations:** School of Mathematics and Statistics, Henan University, Kaifeng 475004, P. R. China; Institute of Applied Mathematics, Henan University, Kaifeng 475004, P. R. China; Laboratory of Data Analysis Technology, Henan University, Kaifeng 475004, P. R. China; School of Mathematics and Statistics, Wuhan University, Wuhan 430070, P. R. China; School of Mathematics and Statistics, Zhongnan University of Economics and Law, Wuhan 430073, P. R. China

**Keywords:** Novel coronavirus pneumonia, Epidemic situation in Henan province, Incubation period, Network analysis, Aggregate outbreak

## Abstract

Based on publicly released data for 1212 patients, we investigated the epidemiological characteristics of COVID-19 in Henan of China. The following findings are obtained: 1) COVID-19 patients in Henan show gender (55% vs 45%) and age (81% aged between 21 and 60) preferences, possible causes were explored; 2) Statistical analysis on 483 patients reveals that the estimated average, mode and median incubation periods are 7.4, 4 and 7 days; Incubation periods of 92% patients were no more than 14 days; 3) The epidemic of COVID-19 in Henan has undergone three stages and showed high correlations with the numbers of patients that recently return from Wuhan; 4) Network analysis on the aggregate outbreak phenomena of COVID-19 revealed that 208 cases were clustering infected, and various people’s Hospital are the main force in treating patients. The related investigations have potential implications for the prevention and control of COVID-19.

## 1 Introduction

Arising from Wuhan South China seafood market at the end of the year 2019, novel coronavirus pneumonia (Named as COVID-19 by WHO since February 11, 2020) has spread for more than one months, and it has directly resulted to more than 74600 confirmed patients, and 2121 people dead till February 20, 2020 [1]-[19]. Various investigations and reports reveal that the COVID-19 is different from the SARS in the year 2003 from the following aspects [1–6] : 1) Higher Infection rate; 2) With incubation period; 3) Incubation period is infectious; 4) Low lethality rate; 5) High mortality among the elderly patients that are with underlying diseases. These features result the prevention and control of COVID-19 to be a difficult task.

As a double-side sword, modern traffic and communication technologies greatly shortage the distances among people and provide convenience for human beings, however, they also facilitate the rapidly and widely spreading of epidemic diseases or rumors [12]. Since January 23, Wuhan has taken strict measures to close the city, and many other places have adopted various measures in succession to prevent the spreading of COVID-19, such as blocking traffic and putting suspicious ones in quarantine. After the outbreak of COVID-19, researchers have paid great efforts on the investigations of COVID-19, including its control and prediction [1]-[18]. Initial investigations pointed out that *bats* are mostly the source of the virus [3], and *Manis pentadactyla* is a potential intermediate host of COVID-19 [11]. The related investigations have great implications on the prevention, control and vaccine development [20]-[30]. Some recent publications have reported the epidemiological and clinical features of patients with COVID-19 [1–6]. To prevent the spreading of COVID-19, the centers for disease control and prevention (CDCs) of many provinces have timely released the data of confirmed cases, which facilitate us to explore the epidemiological and statistical characteristics of patients with COVID-19, and to understand the current situation and characteristics of the epidemic.

As a neighbor of the Hubei province, Henan province is with a large population size, and it is also one of the hardest hit areas of the epidemic. The current total confirmed cases in Henan are only lower than Hubei and Guangdong. The statistical analysis on the patients data in Henan can help us to understand the epidemiological and clinical features of patients [17]. In this paper, based on data collected from CDCs at all levels in Henan, we will perform statistical analysis. Our data includes 1212 patients ranging from January 21 to February 14, 2020, and covering 18 regions of Henan province. We will try to answer the following questions: 1) the current situation and characteristics of the epidemic in the 18 regions of Henan; 2) The sex and age distributions of confirmed cases; 3) The estimation of incubation period of patients with COVID-19; 4) The correlation between the number of patients with Wuhan travel histories and the total number of confirmed cases; 5) Network analysis on aggregate outbreak phenomena. The rest of the paper is organized as follows. Section 2 will introduce our main results and methods; Sections 3 gives some discussions and conclusions of the paper.

## 2 Main Results

### 2.1 The current epidemic situations and characteristics in Henan

From January 21 to February 14, 2020, totally 1212 confirmed cases have been released in Henan. The epidemic situation, time evolution of daily increased infected patients and cumulatively confirmed ones are shown in Fig.1.

**Figure 1.**
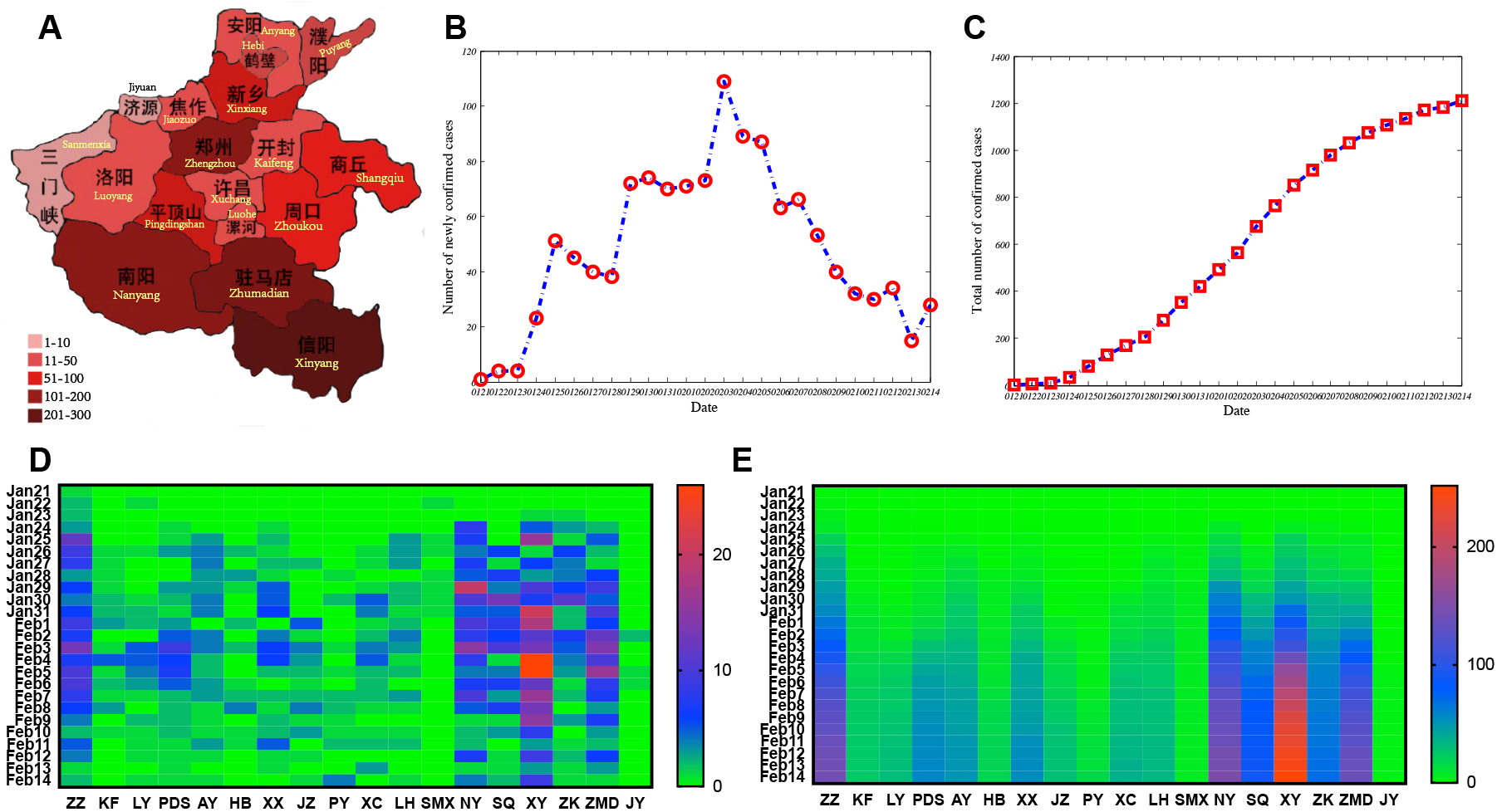
Epidemic situation in Henan till February 14, 2020. A. Epidemic situation in Henan at February 14, 2020; B. The evolution of provincially daily increased confirmed cases; C. The evolution of provincially cumulatively confirmed cases. D. The heatmap of daily increased confirmed cases in 18 regions of Henan province. E. The heatmap of cumulatively confirmed cases in 18 regions of Henan province. Here, ZZ: Zhengzhou; KF: Kaifeng; LY: Luoyang; PDS: Pingdingshan; AY: Anyang; HB: Hebi; XX: Xinxiang; JZ: Jiaozuo; PY: Puyang; XC: Xuchang; LH: Luohe; SMX: Sanmenxia; NY: Nanyang; SQ: Shangqiu; XY: Xinyang; ZK: Zhoukou; ZMD: Zhumadian; JY: Jiyuan. These abbreviations are similarly hereinafter.

Based on Fig.1, we see that the daily increment of patients reaches a peak value of 109 at February 3, 2020. After that, the daily increment of patients significantly decreases. Among the 18 regions of Henan province, Xinyang, Nanyang, Zhumadian, Zhengzhou, Shangqiu and Zhoukou encompass more confirmed cases than the other regions. Xinyang is still with the highest daily increment as compared with the other regions, which is the severely affected area in Henan. In fact, Xinyang, Nanyang and Zhumadian are all near Hubei–the worst affected province in China. It was reported that about five million people have left Wuhan since January 23. According to the investigation from Xu et al. [7, 8], before Wuhan was closed at January 23, Xinyang, Zhengzhou, Nanyang, Zhumadian, Zhoukou, Shangqiu were ranked among the top-50 cities in China that received huge amount of Wuhan personnel. The number of virus carrier is supposed to be proportional to the received amount of Wuhan personnel, which may be one of the main reasons that why the mentioned regions in Henan are so severely affected.

### 2.2 Gender and age distribution

Among the 1212 patients, we abstract the gender information of 1158 patients (95.54% of total samples) and the age information of 1156 patients (95.38% of total samples). From the perspective of both the whole province and its 18 regions, we performed detailed statistical analysis, which are summarized in Fig.2.

**Figure 2.**
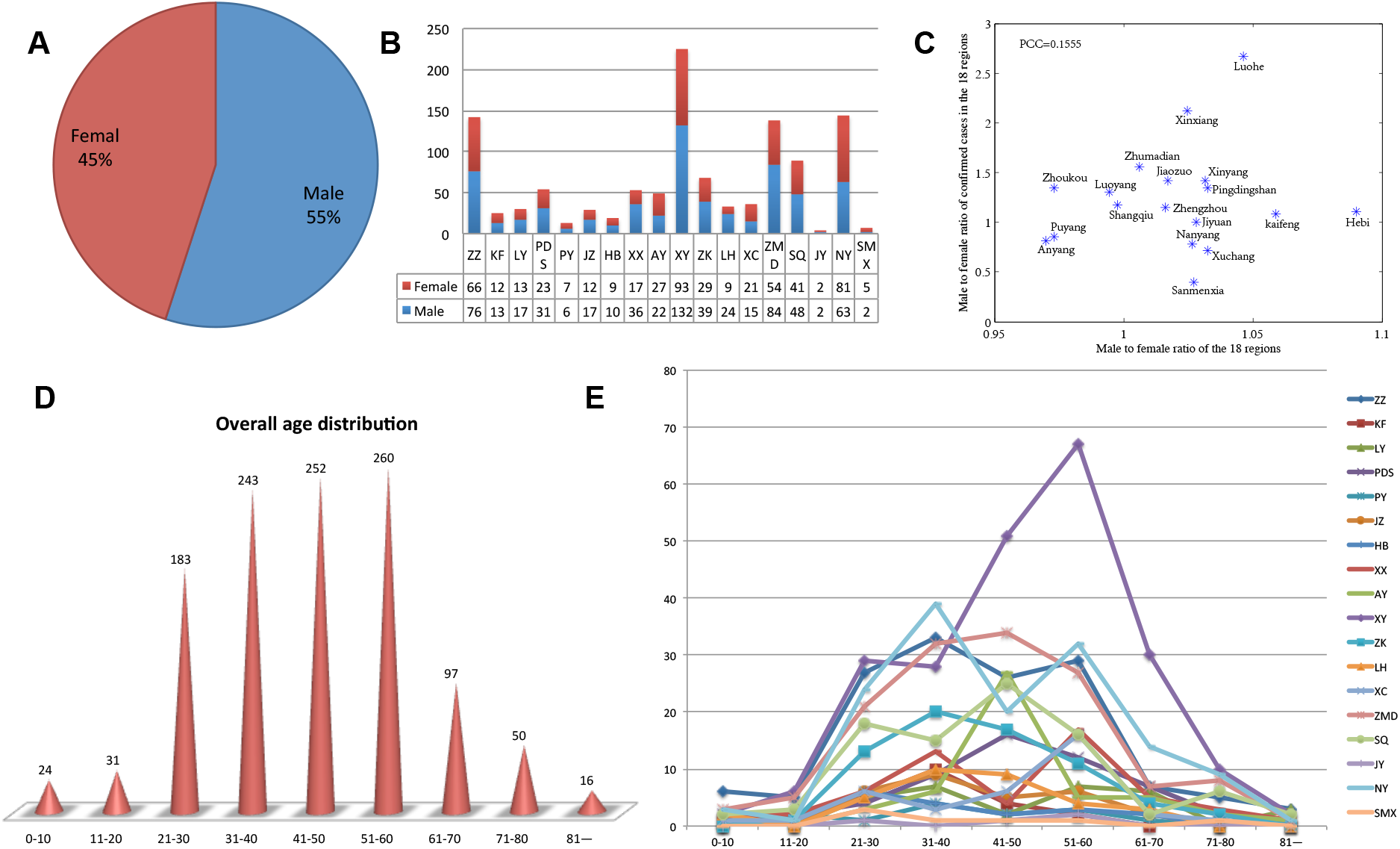
Summary statistics on the gender and age distributions of confirmed cases in Henan. A. MFR in the whole province; B. MFRs in the 18 regions. C. Scatter plot of MFR versus MFPR in the 18 regions of China; D. Age distribution for confirmed cases in the whole province; E. Age distribution for confirmed cases in 18 regions.

Statistical results reveal that 637 out of the 1158 patients are male, which accounts for 55% and apparently higher than females (Fig.2 A), although there are slightly differences among different regions (Fig.2 B). We guess that there are three possible reasons that may result to such gender difference. Firstly, males may be more active and with wider social activities than females, which increases their risk to infect COVID-19. Secondly and may be the most reasonable, an existing medical investigation reported that the expression and distribution of angiotension converting enzyme 2 (ACE2) was wider in male patients than those in females [5]. Similarly to SARS-CoV, it is reported that COVID-19 invades human body through the ACE2 receptor. Based on single cell RNA sequencing technology, researchers have investigated the expression profiles of ACE2 in two male and six female patients with COVID-19 from single cell resolution, and they found that the expression of ACE2 was correlated with gender. The proportion of ACE2 expressed cells is higher in males than in females (1.66% vs. 0.41%) [5]. Besides, the distribution of ACE2 was also wider in males than that in females. Zhao et al. [5] reported that there were at least five different types of cells in the lungs of males with the expression of the ACE2 receptors, while the number was about 2 *∼* 4 in female patients. This may be one of the deep reasons that why male patients were higher than the female ones. Thirdly, one may doubt that the distribution difference may be caused by population structure. To verify whether the male to female ratio (MFR) affects the finding, based on the statistical yearbook report of Henan province in the year 2019, the MFR in the whole province was 1.0126 : 1, which has great difference with the male to female patients ratio (MFPR) (637 : 521 = 1.2226 : 1). Moreover, we perform correlation analysis for the two ratios in the 18 regions, the two ratios show very weak correlation, with Pearson correlation coefficient *r* = 0.1555 (Fig.2 C). Kolmogorov-Smirnov (KS) test [31] on the MFR and MFPR in the 18 regions also shows that the two have significant difference (*KS* = 0.6111, *p* = 0.0018 *<* 0.05). Thus, we speculate that the MFR has no apparent effect on the MFPR. In conclusion, the former two reasons may be the driven force that there was gender difference in confirmed patients.

The age distribution of patients in the whole province or each region all roughly follows normal distribution (Fig.2 D, E). Patients aged between 21 and 60 years old take up more than 81% (938 out of 1156). The main reason may be that people aged between 21 and 60 years old are the social main labor forces, there are many migrant workers in this age interval. Moreover, they may have wider social circles than the others. Therefore, people aged between 21 and 60 years old are key crowd with high risks of COVID-19.

### 2.3 Estimation of incubation period

#### 2.3.1 Definition of incubation period and statistical results

Traditionally, incubation period is defined as the period between the infection of an individual by a pathogen and the manifestation of the illness or disease it causes [32,33]. Different infectious diseases have different incubation periods. However, for a certain infectious disease, its incubation period is relatively fixed. Since the number of pathogens entering the body, virulence and reproductive capacity, as well as power of resistance are different for different people, thus, it is reported that the incubation periods obtained from patients for a certain disease should follow logarithm normal distribution [16,33]. Generally, incubation period can be measured by physiological observations and biological experiments [33]. The determination of incubation period has great implications for disease transmission control and policy making.

The estimation of the incubation period of COVID-19 is a very difficult task. The main difficulty is in that it is difficult to determine the infected time. Based on patients’ information, some works have reported the incubation period of COVID-19. For example, Yang et al. [4] reported that the median incubation period of COVID-19 is 4.75 days; the interquartile range is 3.0 *−* 7.2 days. Hereinafter, we try to estimate the incubation period of patients in Henan. The collected data are with varied quality, many descriptions of patients only told us when the person left Wuhan, or When she/he had suspicious contacts with persons from Wuhan or suspicious persons (Following, we call they are exposed). To facilitate us to estimate the incubation period, we define estimated incubation period as the period from the date of exposure to the date of appearing clinical symptoms or making a definite diagnosis. Apparently, such definition overestimates the actual incubation period. For example, the information from a patient (Mr Zhou, No.2 patients in Nanyang) in Nanyang says that his son left Wuhan and arrived at home at January 6, he and his son were all disease-free for a long period. Until February 3, he became sick and was quickly confirmed with infection of COVID-19. His son was disease-free till now. From our definition of incubation period, his incubation period will be 28 days, which is surprisingly high. We note that, our definition of estimated incubation period may be a little higher than the actually case, since we can not exclude the cases that the person has contacted some other people that carried the COVID-19. That is, we cannot know who was the intermediate spreader (unknown person B), and when she/he transmitted the virus from person A to person C.

Among the collected data, the incubation periods of 483 confirmed patients can be estimated. Statistical results are shown in Fig.3 and Tab.1. The estimated incubation periods for the 483 patients roughly follow the logarithm normal distribution, with a long right tail. Specifically, the 483 confirmed patients were with average estimated incubation period 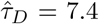days, the mode was 4 days (50 out of the 483 cases) and the median was 7 days (48 out of the 483 cases). About 55% patients were with incubation periods lower than 7 days, while more than 92% patients were lower than 14 days. About 7.45% patients were overestimated with more than 14 days incubation periods.

**Table 1.** Statistical results of estimated as well as fitted incubation period (Fig.3 A) from 483 confirmed patients in Henan. “–”: value is not shown.

| Source | Mean | Mean 95% CI | Median | Mode | Variance | Interquartile range |
| --- | --- | --- | --- | --- | --- | --- |
| Data | 7.4286 | (2.0000,20.0000) | 7.0000 | 4.0000 | 24.1458 | [4.0000,11.0000] |
| ME | 7.4287 | (1.9025,20.1787) | 6.1960 | 4.3103 | 24.1432 | [4.1495,9.3523] |
| OLS | 10.6840 | (1.7561,35.9274) | 7.9431 | 4.3903 | 92.3723 | [4.7254,13.3519] |
| MLE | 9.1463 | (1.2202,33.5482) | 6.3981 | 3.1308 | 87.3000 | [3.6175,11.3160] |

**Figure 3.**
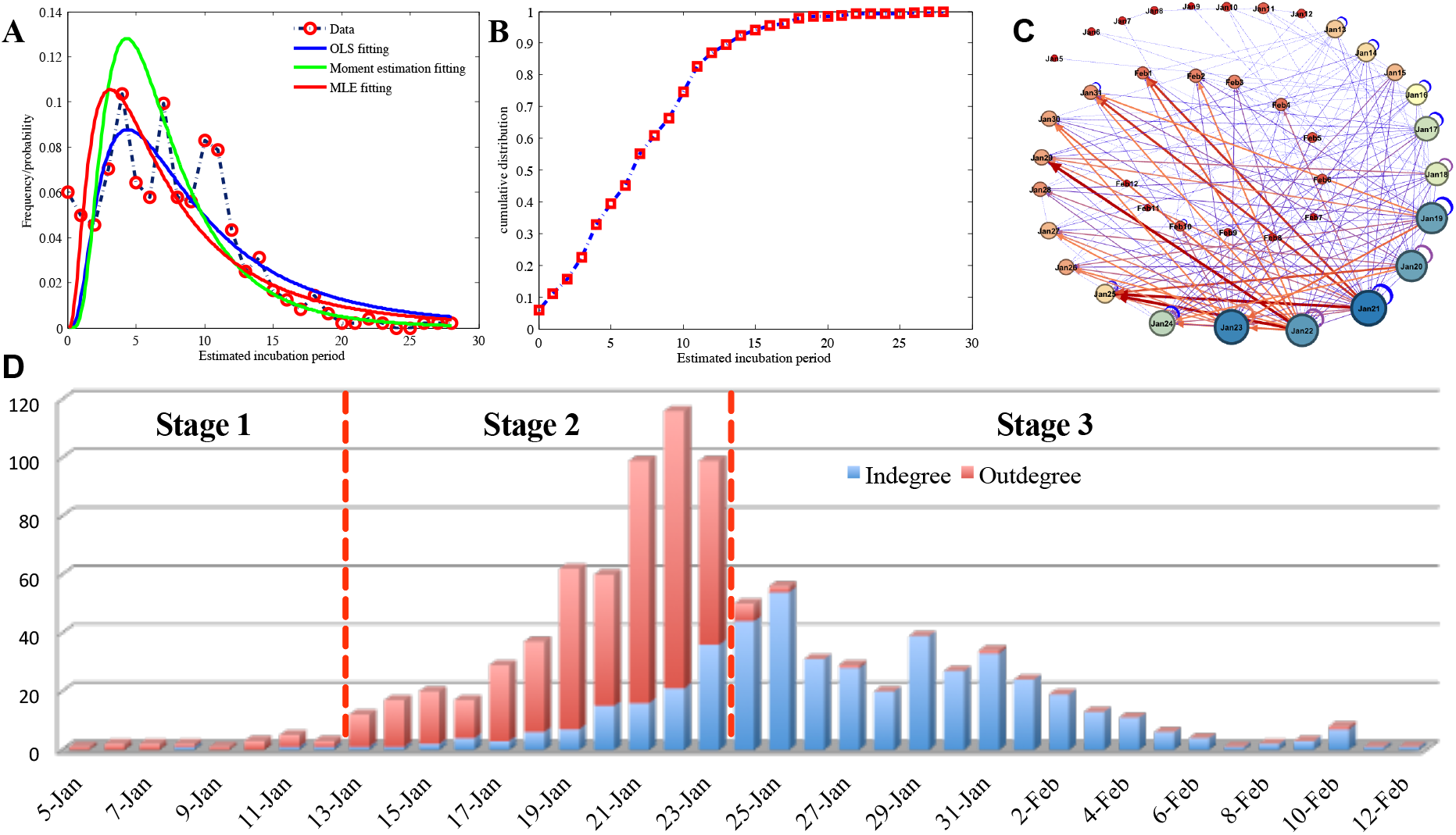
Statistical results of estimated incubation periods for 483 confirmed patients in Henan province. A. Frequency distribution of estimated incubation periods and the probability density curve for fitted incubation period (We consider moment estimation (ME), maximum likelihood estimation (MLE) and ordinary least square (OLS) estimation of parameters). B. The cumulative frequency distribution of estimated incubation periods for the 483 patients. C. Transfer diagram from exposure to infection for the 483 confirmed patients. Nodes at the two ends of an edge correspond to the exposed date and the date with clinical symptoms or confirmed infected. Edges represent the transfer from the two dates, and thicknesses of edges are proportional to the numbers of patients. Self-loops mean that the dates for exposure and appearing clinical symptoms or confirmed infected were the same. D. Weighted indegree and outdegree distributions of the directed graph that was shown in C. Weighted indegree denotes the total number of patients with clinical symptoms or confirmed infected; while weighted outdegree represents the total number of exposed persons that will be confirmed to be infected later.

Due to the definition, the estimated incubation periods in this paper may be longer than the real cases. However, the distribution has some implications. It indicates that a few patients may be with very long incubation period, which increase the difficulty to control and prevent the COVID-19.

#### 2.3.2 Modeling the distribution of the incubation period

To theoretically model the incubation period, we suppose the incubation period *τ* follows the logarithm normal distribution [16, 33]:

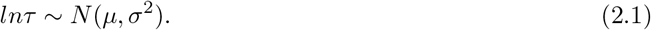

That is, the probability density function (PDF) of *τ* can be written as

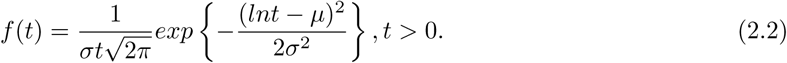

Here, *µ, σ*^2^ are two parameters that can be estimated from data. We consider three different approaches to estimate the parameters *µ* and *σ*^2^, including the moment estimation (ME), the ordinary least square (OLS) estimation and the maximum likelihood estimation (MLE) [34]. As to the ME, based on the PDF of incubation period *τ*, we can easily obtain

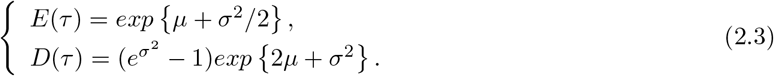

Setting *E*(*τ*) and *D*(*τ*) with the estimated incubation period from the 483 patients (shown in Tab.1), we obtain the following equations.

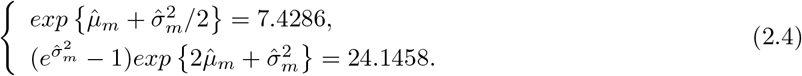

Using the fsolve function (which is based on the OLS method) in Matlab, we obtain 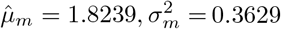The fitted PDF curve can be found in Fig.3 A, and some theoretical estimations of the incubation period based on the ME method are summarized in Tab.1.

Similarly, for the OLS estimation, *µ* and *σ*^2^ can be estimated from the following optimization objective function:

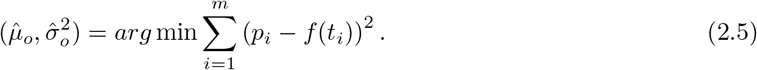

Here, *p*_*i*_ = *n*_*i*_*/n* denotes the ratio of patients with incubation period *i*(*i* = 0, 1, …, *m*) (totally *m*+1 unique incubation periods), which can be easily computed from the 483 patients (n=483). *f* (*t*_*i*_) corresponds to the PDF with incubation period *t*_*i*_, which is a function of the unknown parameters *µ* and *σ*^2^. Since *τ /*= 0 in Eq.(2.2), we omit the case *τ* = 0 (only 29 cases were with *τ* = 0). By solving the optimization problem (2.5), we obtain 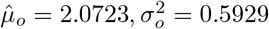

For the MLE, the estimation of *µ* and *σ*^2^ can be obtained from minimizing the following negative logarithmic likelihood function.

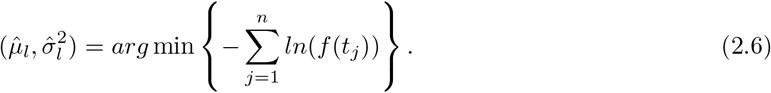

Here, *t*_*j*_(*j* = 1, 2, …, *n*) denotes the incubation period for the *j*’th patient. Similarly to OLS, the case with *τ* = 0 will be not considered. Finally, we obtain

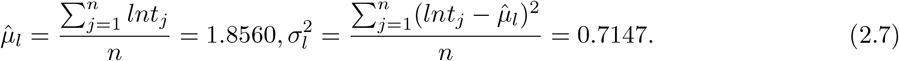

The fitted curves from the three methods are shown in Fig.3 A. Comparing among the three methods, it seems that the fitted curve from the ME method has the smallest variance, the variances of the OLS and MLE approaches are both very high. High variance indicates higher proportions of patients with long incubation periods. The ME method may well mimic the actual long right tail of the incubation period, while the OLS estimation could well describe the cases with middle incubation periods (incubation periods 2 *∼* 14). The MLE approach seems in between the ME and OLS approaches. The estimated interquartile range from the ME method is [4.1495, 9.3523], which closes to the result from the 483 patients ([4,11]).

The theoretical modeling of incubation period further illustrates that the incubation period may follow the logarithmic normal distribution. Moreover, it can be used to estimate the actual statics of incubation periods, and provide valuable references for decision-making.

#### 2.3.3 Transfer diagram from exposure to infection

The epidemic of COVID-19 in Henan has undergone three stages (Fig.3 C and D). For the first stage that before January 13, both exposed and confirmed patients were not so much, the number of exposed (outdegree) ones were more than the confirmed ones (indegree). While for the second stage that between January 13 and January 23, the COVID-19 was in outbreak period, both exposed and confirmed ones were considerable, and the number of exposed ones are far more than the confirmed ones. During this period, many college students and migrant workers in Wuhan return their hometown, the large number of people flow from Wuhan increased the risks of exposure and infection. After January 23, the epidemic entered the third stage, the number of daily confirmed cases were apparently higher than exposed ones, and the confirmed cases roughly decreased with time. This may be attributed to the adoption of prevention and control measures. With the closing of the Wuhan city at January 23, many regions of Henan quickly adopt very strict measures to prevent the spreading of COVID-19. Thus, after January 23, the number of exposed ones became fewer and fewer (Fig.3 D), which is a good phenomena, indicating that the prevention and control measures become to gradually at work.

### 2.4 Correlation with Wuhan travel histories

For the 18 regions of Henan province, we also summarized the numbers of currently confirmed patients with Wuhan travel histories (denoted as vector *X*_1_), the numbers of patients without Wuhan travel histories (*X*_2_), the numbers of patients that have contacts with persons from Wuhan (*X*_3_), as well as the numbers of patients that are without Wuhan travel histories, but who have contacts with suspicious persons (*X*_4_). The barplots for the numbers of the four categories of patients in the 18 regions are shown in Fig.4 A (In total of 1149 patients were known with or without Wuhan travel histories). Among patients with Wuhan travel histories, the released information of 250 patients contained transportation information. We found that 46% of the 250 patients have recently traveled by train or by bus, these patients may transmit the COVID-19 to a lot of people that in the same train or bus.

**Figure 4.**
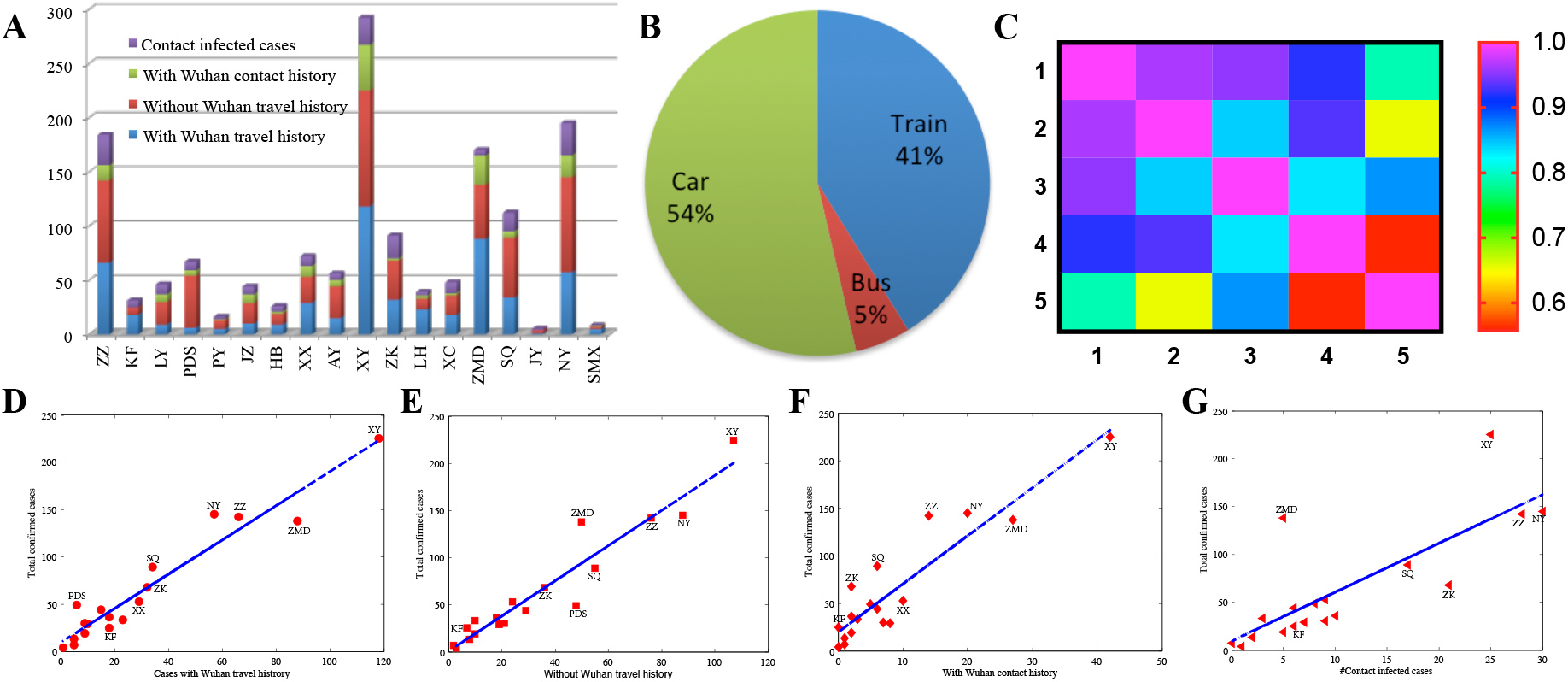
Correlation between COVID-19 infection and Wuhan travel histories. A. Bar-plots for the numbers of four categories of patients in the 18 regions of Henan province. B. Transportation manners of 250 patients that came from Wuhan. C. The heatmap of the correlation matrix among *Y, X*_1_ *−X*_4_ (corresponding to 1-5 respectively). D-G. The scatter plots of *Y* versus *X*_1_ *− X −* 4 respectively and the corresponding fitted linear regression lines.

For the 18 regions, we investigate the correlation between the currently total numbers of confirmed patients (*Y*) with *X*_1_ *− X*_4_ respectively. The heatmap of the correlation matrix is shown in Fig.4C. It reveals that *Y* has the highest correlation with *X*_1_, the Pearson correlation coefficient between the two is 0.9643, the fitted regression equation [35] is

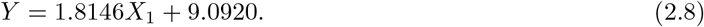

Such linear relationship statistically holds (*F* = 212.31, *p <* 0.0001, *R*^2^ = 0.9299). However, though there are certain correlation between *Y* and *X*_2_ *− X*_4_, their strength of correlations are all lower than that between *Y* and *X*_1_ (Fig.4 D-G). This indicates that persons with Wuhan travel histories have high risks of infection with COVID-19, and regions with more such persons tend to encompass more patients infected with COVID-19.

### 2.5 Network analysis on aggregate outbreak phenomena

From the 1212 patients, we constructed a heterogeneous network and performed some analysis on such network (Fig.5). The network contains 1105 patients that have been treated in 248 hospitals, 123 inter-hospital transfer relationships that involved 206 patients, and 208 patients that were clustering infected. Nodes in the network include patients and hospitals. Edges among patients represent relationship between relatives, friends or colleagues; Edges between patients and hospitals denote the patients were treated in the related hospitals; while edges among hospitals indicate the inter-hospital transfer treatments. Sizes of nodes were proportional to nodes’ degree. Thickness of edges was proportional to the numbers of patients.

**Figure 5.**
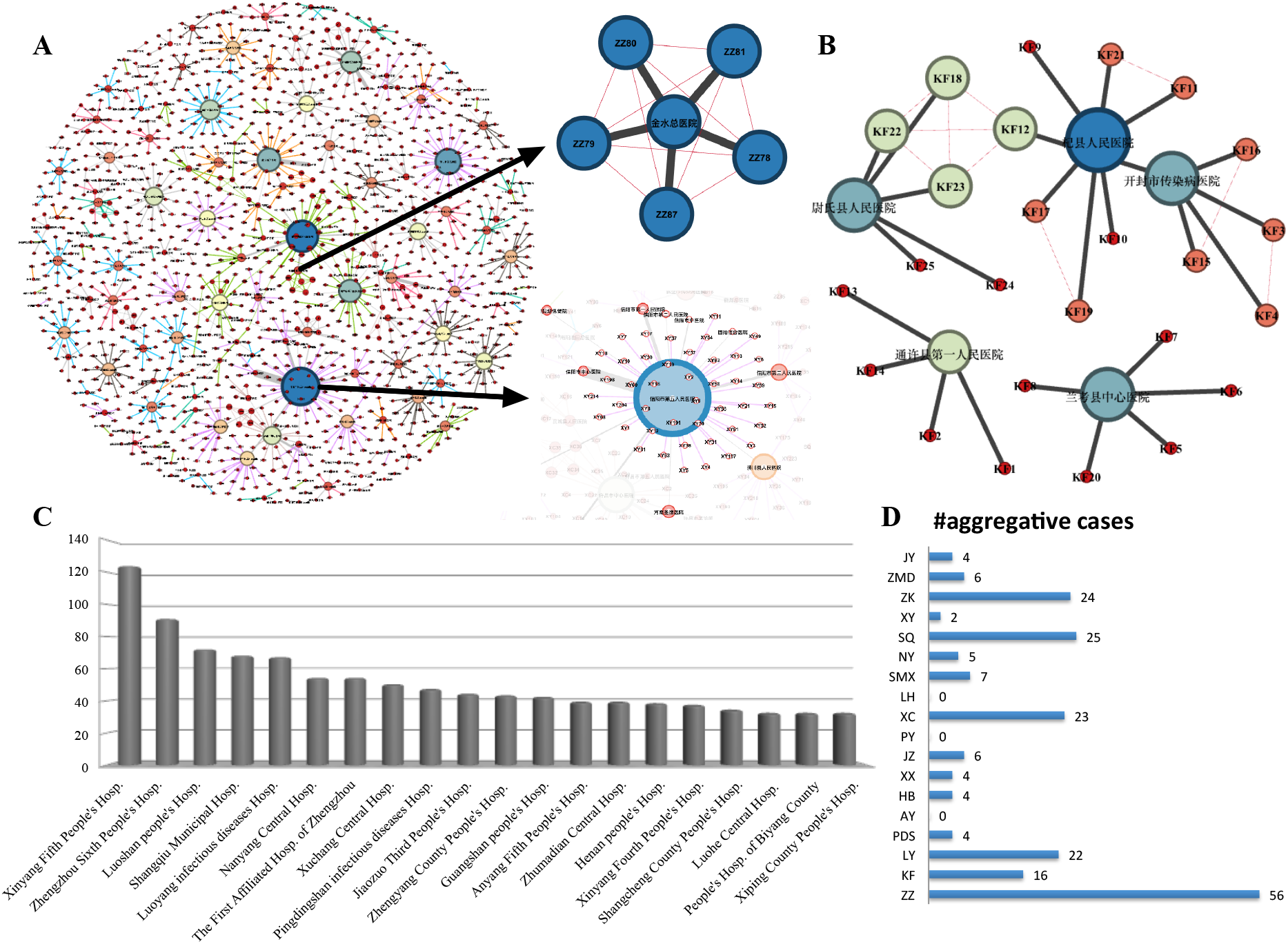
Network analysis among patients, hospitals and their relationships. A. A heterogeneous network that contains 1105 patients, 248 hospitals, 123 inter-hospital transfer relationships that involved 206 patients and 208 clustering infected patients. B. The heterogeneous network for patients in Kaifeng. C. The top-20 hospitals that are with the most patients in treatments. D. Distributions of the 208 aggregate outbreak patients in the 18 regions of Henan.

From Fig.5, we see that a few hospitals encompass a large number of patients in treatments, and the aggregate outbreak phenomena were ubiquitous. For example, the five patients that are treated in Zhengzhou Jinshui General Hospital are relatives to each other, they were successively infected. Similarly, 12 patients in Kaifeng are also relatives to each other. In fact, totally 208 patients were clustering infected, and Zhengzhou, Shangqiu, Zhoukou, Xuchang and Luoyang encompassed the most aggregative outbreak patients. Among the designated hospitals in Henan province, people’s hospitals in different cities encompass a great number of patients. Xinyang Fifth People’s Hospital received the most patients in Henan, followed by the Zhengzhou Sixth People’s Hospital and the Luoshan County People’s Hospital. Network analysis can provide us important information about patients, hospitals and their relationships; it can also provide valuable guidance for the distribution of epidemic prevention materials.

## 3 Discussions and conclusions

Through collection of publicly released patients information by various CDCs in Henan province during January 21 to February 14 of 2020, we performed statistical analysis on 1212 confirmed patients infected with COVID-19. The number of newly confirmed cases reached its peak at February 3, and it started to fall after that day. Among the 18 regions of Henan, Xinyang is with the heaviest epidemic, and it is currently also with the highest daily increment. We found that 55% patients in Henan were male, which is apparently higher than the females. We found that the gender difference of patients in Henan is 1.2226 : 1, while from Yang et al. [4], they reported that the gender difference for 4021 patients from over 30 provinces is 0.31 : 0.27 = 1.1481 : 1. Our result is close to the existing ones. We reported that the MFRs of the whole province and its 18 regions are not the main factor that results to the gender difference in patients. Two possible main factors that result to such gender difference of patients include the expression and distribution of ACE2s were wider in male patients than those in females, and 2) males are more active and with wider social activities than females in many regions of Henan. The ages of patients generally follow normal distribution, and with more than 81% patients are aged between 21 and 60 years old.

Incubation period is very important but very difficult to be estimated, since it is difficult to know when the patients were infected. We define the period from firstly contacting with suspicious persons or recently return from Wuhan to firstly appearing clinical symptoms (fever, cough etc) as incubation period, and we estimate its distribution. Statistical analysis on 483 patients reveals that the average estimated incubation period was 7.4 days, the mode was 4 days. About 55% patients were with incubation periods lower than 7 days, while more than 92% patients were lower than 14 days. About 7.45% patients were overestimated with more than 14 days incubation periods. Based on ME, OLS and MLE approaches, we modeled the incubation period as logarithmic normal distributions, and we found the ME method can well mimic the long right tails of the PDF for the incubation periods. Based on the ME approach, interquartile range of theoretically estimated incubation period is between 4.1 and 9.4 days. It is noted that, due to our definition of incubation period, the estimated values are unavoidably higher than the actual cases, however, the distribution and statistics from the estimated incubation periods are undoubtedly meaningful for real-world decision-making.

We also find that the numbers of patients with recent Wuhan travel histories are highly correlated with the total numbers of confirmed cases in the 18 regions of Henan. During our analysis, we consider the linear relationship between *Y* with each *X*_*i*_, which mainly for the purpose of avoiding multicollinearity [34] among *X*_*i*_(*i* = 1, 2, 3, 4). Furthermore, we perform network analysis on aggregate outbreak phenomena of COVID-19 in Henan, we reported that 208 cases were clustering infected ones. Various people’s Hospital in Henan are the main forces in treating patients. The related investigations may provide valuable suggestions and guidance for the prevention and control of COVID-19.

## Data Availability

Data are publicly available from the all levels of CDCs' websites in Henan of China

## Acknowledgements

This work was supported by the National Natural Science Foundation of China (Grant Nos. 61773153, 61773175, 61573262).The Supporting Plan for Scientific and Technological Innovative Talents in Universities of Henan Province (Grant No. 20HASTIT025), and the Training Plan of Young Key Teachers in Colleges and Universities of Henan Province (Grant No. 2018GGJS021).Partly supported by the supporting grant of Bioinformatics Center of Henan University (Grant No. 2018YLJC03). We thank the valuable discussion with associate professor Shaoli Wang and Aimin Chen at Henan University. We also want to express our grateful for the valuable comments and feedback from the readers of the first version of the paper, which was released on the WeChat official accounts of the Swarma Club at February 13, 2020.

## Notes

### Competing Interest Statement

The authors have declared no competing interest.

### Clinical Trial

This is an epidemiological study on patients with COVID-19 in Henan, China

